# *Echinacea* as a Potential Force against Coronavirus Infections? A Mini-Review of Randomized Controlled Trials in Adults and Children

**DOI:** 10.1101/2021.12.23.21267893

**Authors:** Nicolussi Simon, Ardjomand-Woelkart Karin, Stange Rainer, Gancitano Giuseppe, Klein Peter, Ogal Mercedes

## Abstract

*Echinacea purpurea* was shown to broadly inhibit coronaviruses and SARS-CoV-2 in vitro. This review discusses the available clinical evidence from randomized, blinded and controlled human studies. Two RCTs with results on enveloped viruses, respectively coronavirus infections during prevention treatment were detected. Incidence and/or viral loads were measured by RT-PCR and symptom severity was recorded. Jawad et al. (2012) collected nasopharyngeal swabs from adults (N=755) over 4 months of continuous prevention. Overall, 24 and 47 enveloped virus infections occurred, including 21 and 33 coronavirus detections [229E; HKU1; OC43] with Echinaforce® extract [2’400mg daily] and placebo, respectively (p=0.0114). Ogal et al. (2021) administered the same extract [1’200mg] or control for 4 months to children (4 – 12 years) (N=203). *Echinacea* reduced the incidence of enveloped virus infections from 47 to 29 (p=0.0038) whereas 11 and 13 coronavirus detections [229E, OC43, NL63] were counted (p>0.05). Respiratory symptoms during coronavirus infections were significantly lower with area-under-curve AUC=75.8 (+/-50.24) versus 27.1 (+/-21.27) score points (p=0.0036). Importantly, viral loads in nasal secretions were significantly reduced by 98.5%, with Ct-values 31.1 [95% CI 26.3; 35.9] versus 25.0 [95% CI 20.5; 29.5] (p = 0.0479). Results from clinical studies confirm the antiviral activity found for *Echinacea* in vitro, embracing enveloped respiratory pathogens and therefore coronaviruses as well. Substantiating results from a new completed study seems to extrapolate these effects to the prevention of SARS-CoV-2 infection. As hypothesized, the testified broad antiviral activity of *Echinacea* extract appears to be inclusive for SARS-CoV-2.

## 1. Introduction

Zoonosis describes the transmission of pathogens from animals to humans, which is not a new phenomenon by itself. Earlier events of zoonosis included the transmission of yersinia pestis from rodent flea (plague) or Ebola spilling over from apes causing millions of deaths in Europe and Africa. Respiratory viruses like influenza were repetitively seen crossing species barriers from animals to humans triggering deadly disease outbreaks like Spanish (H1N1), Asia (H2N2) or Hong Kong (H3N2) flu epidemic as well as the more recent bird flu caused by avian H5N1 virus [1].

The 21st century has seen another zoonotic thread, coronaviruses, with successfully contained outbreaks of SARS-CoV in 2002/2003 or Middle East Respiratory Syndrome (MERS-CoV) occurring from 2012 to 2015 [2]. Most coronaviruses could be traced back to be of animal origin whereas camels, bats or civet cats are likely natural hosts or vectors [3,4]. Evidence exists that even the Russian flu was not caused by influenza but by coronavirus strain OC43, due to its highly similar symptomatology to current Covid-19 [5]. Notably, most endemic coronaviruses we know today, originally spilled over from other species as reviewed elsewhere [6]. Due to their high pathogenicity and rather short incubation period, previous coronavirus outbreaks were relatively easy to curb and thus limited to restricted areas. In contrast, SARS-CoV-2 is much trickier to contain due to its ability to disseminate through “silent transmitters” during the extended incubation period of up to 14 days [7]. Since the newly emerging SARS-CoV-2 was classified as a high-risk pathogen on February 29th 2020 reaching pandemic status in March 11th 2020 the world’s endeavors strive to control the dissemination of the virus [8].

Vaccines are a highly efficient means for disease prevention in general and their development nowadays has tremendously been shortened. Nevertheless, from first occurrence (patient zero), virus isolation and determination to large-scale production, distribution of vaccines and final attainment of herd immunity valuable time passes. As the pandemic lingers on, new SARS-CoV-2 variants emerge through inaccurate genetic replication with altered biological behavior and fitness. Several of those variants-of-concern (VOC’s) have meanwhile been discovered which display in comparison to the original WUHAN strain more severe pathogenicity and immune evasion potential (e.g. delta variant B.1.671.2 or Omicron B.1.1.529) [9–12].

Because the preventive potential of vaccination is never complete and depending on circulating variants, the time point of immunization or an individual’s immune status, the demand for novel antivirals for containment of coronavirus infections remains high [13]. Some preparations and practices claim to bolster the immune defense to keep at bay and overcome any potential viral infection. Despite lacking proof of effectiveness for the particular pathogen these preparations are intensely consumed.

In 2020 scientists have speculated whether usage of minerals, vitamins or herbal preparations would be of any help for preventing or treating Covid-19 illness, but recommendations mostly relied on theoretical considerations, epidemiological observations, in vitro experiments rather than appropriately controlled intervention trials and therefore remained speculative [14–17]. *Echinacea*, known as the immune herb was highly demanded during the current pandemic and sales steeply increased from 2019 to 2020 by 36.8% in the US [18]. In 2020, Signer et al. published in vitro data revealing a broad virucidal activity for *Echinacea purpurea* (hydroethanolic extract (65% v/v) of freshly harvested *Echinacea purpurea* herb and roots manufactured in GMP quality) against all kinds of coronaviruses ranging from the typical common cold CoV-229E to highly pathogenic SARS-CoV-2 viruses [19]. Clinical relevance of findings however remained uncertain, because direct contact of pathogen with the extract was prerequisite for full activity, which was thought to be the case upon sucking tablets/gargling tincture in the pharynx.

Besides being antiviral, *Echinacea* influences the immune system in a manner best described as adaptive immune-modulation, rather than pure immune stimulation. *Echinacea* significantly reduced the inflammatory cytokines TNF-alpha and IL-1-beta by up to 24% compared to baseline and increased the anti-inflammatory cytokine IL-10. In addition, there was an increase of up to 50% IFN-γ [20,21]. Further immunomodulatory mechanisms of *Echinacea* were shown to involve potent activation of the endocannabinoid system (ECS) by specific alkylamides via the CB2 receptor [22,23]. Several of these bioactive N-alkylamide principles are structurally similar to endocannabinoids (e.g. anandamide) which influence the cytokine milieu in human whole blood at low nanomolar concentrations [24]. During COVID-19 progression, activation of the endocannabinoid system (ECS) could be an additional approach against systemic inflammation and the cytokine storm [25]. *Echinacea* could therefore reduce inflammatory processes through synergistic immunopharmacological targeting of CB-receptors, mild inhibition of the fatty acid amide hydrolase (FAAH) or endocannabinoid transport [26,27]. In addition to immunoinflammatory changes, during viral infections, macrophages and neutrophils can produce numerous ROS including hydrogen peroxide (H2O2), superoxide (O2•-) and hydroxyl (OH•) radicals, which further activate several signaling pathways and institute inflammation and cell death in multiple organs, including the lungs [28].

Preclinical studies thus attribute pharmacological actions to *Echinacea*, which could be beneficial for the prevention and treatment of viral diseases. The goal of this review was to identify coronavirus infections in clinical studies on *Echinacea* and to evaluate preventive and treatment benefits.

### 2. Materials and Methods

We searched the available literature on MEDLINE and EMBASE from inception up to 23th March 2021 limited to human studies and included all synonyms of the following MeSH terms: *Echinacea* species OR coronavirus OR virus infections AND Roter Sonnenhut or Purple Coneflower OR *Echinacea purpurea* OR human clinical studies OR respiratory tract infections AND terms related to study design (RCTs). A detailed search strategy is shown in Appendix 1.

Obtained articles were further selected for clinical trials studying respiratory tract infections in humans and detection of respiratory viruses independently by SN and GG. In case of discrepancy regarding eligibility of articles, consensus was sought between all authors prior to inclusion or rejection. The research outcome was to be presented in a flow chart as requested by PRISMA [29]. In case of inconsistencies or missing information, the corresponding authors of the published article was contacted for further information. European and US clinical trials registers, EudraCT and clinicaltrials.gov were screened for clinical studies on *Echinacea* and respiratory tract infections and researchers contacted for possible results.

Data extraction was undertaken using a pre-defined form retrieving the following information: primary author’s name, publication date, country, study design, sample size, researched plant species and preparation, duration of treatment (prevention vs acute treatment), dosage, incidence of coronavirus virus infections and virus concentration (Ct-values), information regarding symptomatic development (e.g., total symptom score, episode duration, area-under-curve or similar). The methodological quality of included studies was assessed using the criteria as proposed by Jadad [30]. Results are provided for safety (SAF), intention-to-treat (ITT) or as indicated for the per protocol (PP) populations.

### 3. Results

A total of N=1687 articles were identified through database searching including PubMed and EMBASE using a combination of keywords, which includes Medical Subject Headings (MeSH)/Emtree terms (Embase Subject Headings). For further details see Appendix 1.

N=988 articles were excluded at the initial screening due to duplicated and irrelevant articles. Sixty potentially relevant articles were selected for the full text assessment, of which 58 were further excluded due to studies that were not reporting respiratory tract -or coronavirus infections, *Echinacea* species, non-RCT and human studies (n= 57) or combination of Zingiber officinale and *Echinacea* (N=1), which is detailed in PRISMA flow chart (Figure 1). Finally, two studies selected for the systematic review, which met the eligibility criteria of the current study (Table 1).

**Table 1.** Included studies with details on methodology and outcome.

| Author | Population | Country | Jadad Score | Plant species | Preparation | Prevention Dosis | Comparator | Sampling source | Analysis method | Symptomatic Assessments |
| --- | --- | --- | --- | --- | --- | --- | --- | --- | --- | --- |
| Jawad M, et al. 2012 | Adults > 18 years, N=755 | UK | 5 | *Echinacea purpurea* | Alcoholic extract of *Echinacea purpurea* herb/roots | 3 x 20 drops /d | Placebo | Nasopharynx | RT-PCR, no Ct-values reported | Individual and total symptom scores, Area-under-curve |
| Ogal M, et al. 2021 | Children 4 – 12 years, N=203 | CH | 4 | *Echinacea purpurea* | Echinacea Junior tablets, *Echinacea purpurea* herb/roots | 3 x 1t ablet /d | Vitamin C | Nasopharynx | RT-PCR, Ct-values reported (semi-quantitative) | Individual and total symptom scores, Area-under-curve |

**Figure 1:**
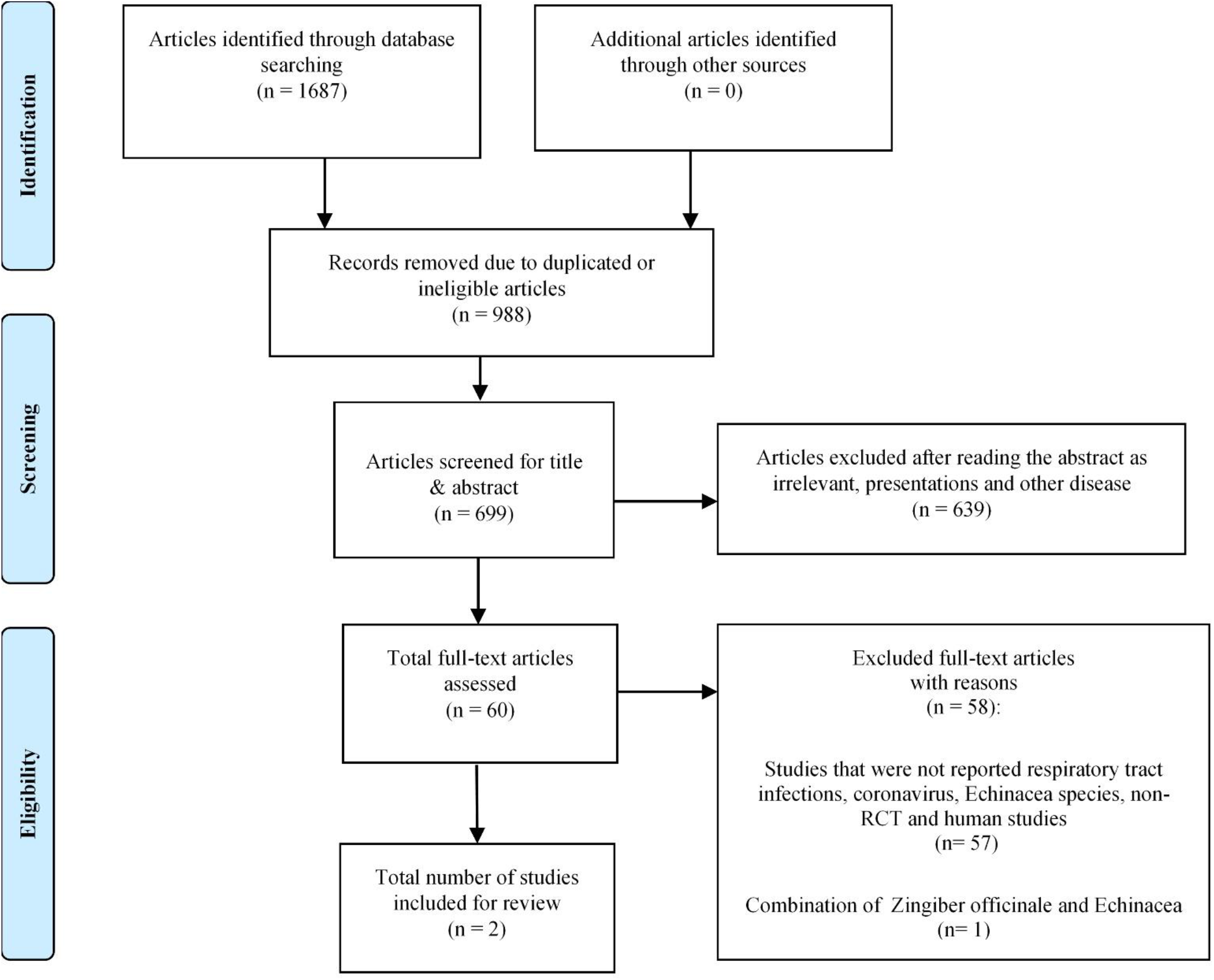
PRISMA flow chart for systematic reviews detailing the database searches, the number of publications screened and eligible results retrieved.

Due to the low number of referenced studies we abstained from carrying out a quantitative meta-analysis. Instead, we decided to qualitatively discuss the observed findings from the two contributions, which are detailed in the following.

A first clinical study was carried out by Jawad et al. at Cardiff University, United Kingdom as double-blinded placebo-controlled monocentric RCT. Over a period of 4 months, participants from Cardiff, Wales, UK over 18 years were to apply a solution of 3x 0.9mL daily. The hydro-ethanolic extract (57.3 % m/m) from freshly harvested aerial parts of *Echinacea purpurea* to 95 % supplemented with 5 % *Echinacea purpurea* roots extract was used from a commercial preparation (Echinaforce® drops, A.Vogel AG, Switzerland, licensed there as phytodrug), standardized to contain 5 mg/100 g of dodecatetraenoic acid isobutylamide, based on high performance liquid chromatography measurements. It corresponded to the standard adult dosage of 2’400 mg/d of the spissum extract from the same manufacturer, of which 1’200 mg/d were used as recommended dosage for children in the second trial. The placebo solution was similar in shape, color, consistency, odor, flavor and amount of alcohol [31]. Patients were advised to keep the remedy in the mouth for 10 seconds prior to swallowing. The study looked at the occurrence of cold days and respiratory tract episodes as defined by Jackson et al. and assessed symptoms via diary [32]. Alongside symptom recording, patients were to take one nasopharyngeal sample on day 3 of each episode for identification of the respiratory pathogen using xTag FAST Track Multiplex Panel (Luminex, Austin Texas). Analysis was carried out by the Provincial Health Services Authorities, PHSA; BC Center for Disease Control, Vancouver, Canada. The following pathogens were screened: Influenza A H1/H3, Influenza B, Respiratory Syncytial Virus, Coronavirus 229E/OC43/NL63/HKU1, Parainfluenza virus 1–4, human Metapneumovirus, Entero-rhinovirus, Adenovirus, and human Bocavirus. The methodological quality according to Jadad et al. reached the maximum score of 5.

Overall, N=755 subjects were randomized to E. purpurea or placebo and N=717 subjects were eligible for analysis as per safety collective (SAF). Preventive benefits were reported on the level of total cold episodes, cumulative cold days and recurrent episodes. The *Echinacea* group (N=355) provided 86 nasopharyngeal samples in comparison to 115 samples from control group (N=362) of which 54, resp. 74 samples tested positive for any respiratory virus (p=0.0663). In 24, resp. 47 samples the presence of enveloped viruses was verified, and the resulting odds ratio OR = 0.49 calculated to be statistically significant (p = 0.0114, X2 test). The original publication however did not provide further information on coronavirus infections, which could be retrieved by correspondence from authors of this study: In the *Echinacea* group 9 detections of CoV-229E, 11 of HKU1 and 1 of OC43 cumulated to overall 21 coronavirus infections (overall incidence rate 5.9%) in comparison to 15 detections of CoV-229E, 17 with HKU1 and 1 with OC43 accruing to 33 infections for placebo (overall incidence rate 9.1%). The resulting odds of contracting infection was as OR = 0.63 similar to the overall results on cumulated enveloped viruses (OR = 0.49; 0.0114, X2 test) (Figure 2). Preventive effects on coronaviruses were statistically significant in a collective of participants (*Echinacea* N=163 and placebo N=165), who have actively utilized their patient diary to report adverse events and/or cold-related symptoms (ITT sub-group analysis [33]). Here, an overall incidence rate of 5.5% for *Echinacea* opposed a rate of 14.6% for placebo (p = 0.010, X2 test). Ct values of individual virus detections were not analyzed in this study and therefore conclusions regarding viral loads not possible. Although fewer coronavirus infections were detected with *Echinacea* extract, the mean duration of episodes was with 5.8 days [95% CI: 4.4-7.2] versus 5.9 days [95% CI: 4.4-7.5] unchanged.

**Figure 2.**
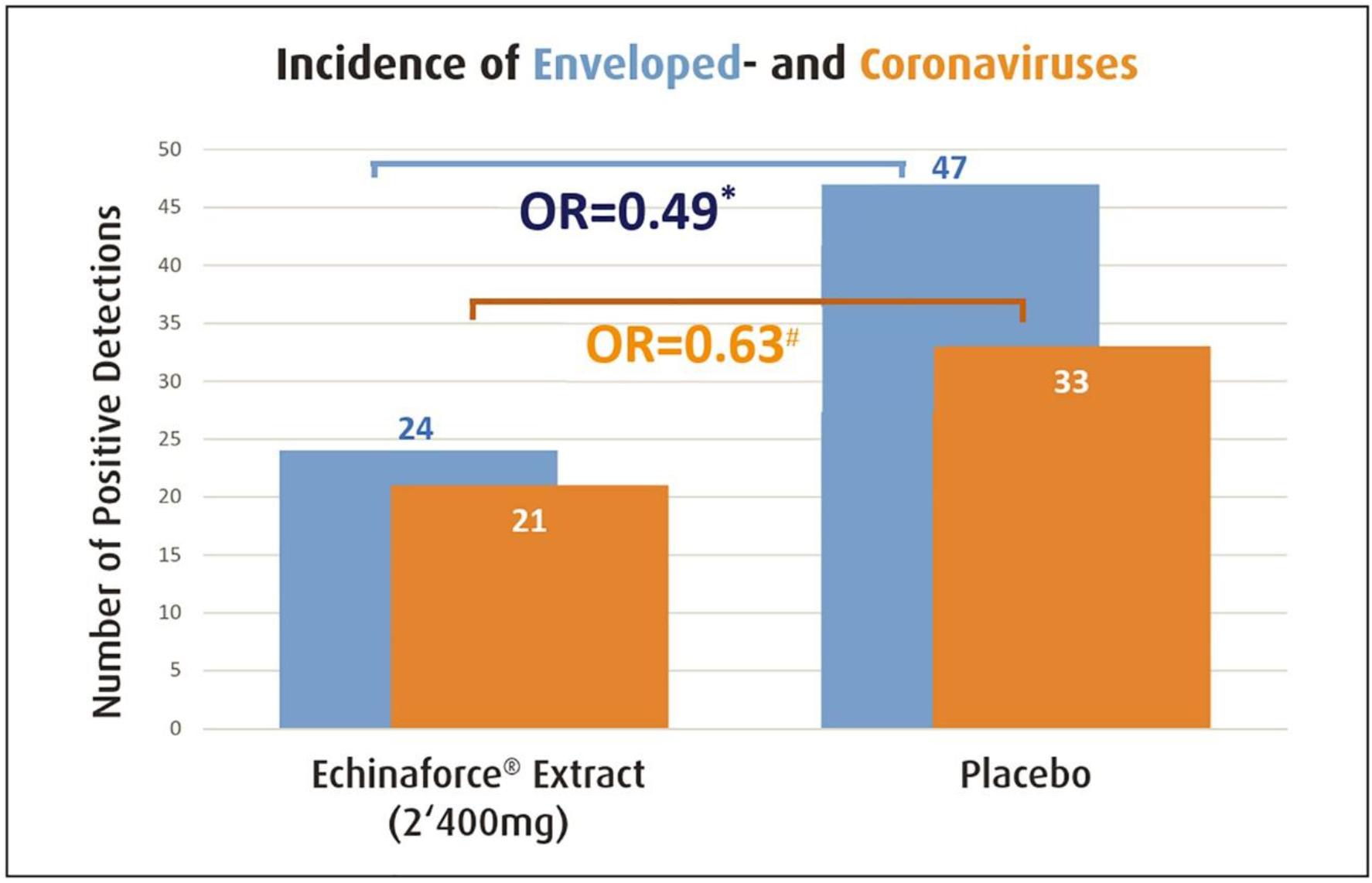
*Echinacea* significantly prevented enveloped virus infections (blue bar, * p=0.0114) and thereof coronaviruses 229E, HKU, NL63 and OC43 (orange bar, ♯ p>0.05) as measured by Jawad et al. 2012. Prevention of coronaviruses was significant for the ITT sub-group (p=0.010).

In a second double-blinded multi-center RCT, the same *Echinacea* extract formulated in tablets was given to children 4 to 12 years old living in central Switzerland [34]. Here, the authors chose low-dose vitamin C 3x 50 mg/d as control. The commercial product was the child-friendly *Echinacea* formulation (Echinaforce® Junior tablets, same manufacturer A. Vogel AG, Switzerland, also licensed there as phytodrug) with a recommended dose of 1,200 mg/d for children over 4 years. This trial enrolled 203 principally healthy children in winter season 2016/17, of which N=103 were randomized to Echinacea 3x 400 mg/d and N=98 to Vitamin C 3×50 mg/d. The preparations were administered over a period of 4 months including a 1-week treatment break after 2 months. Parents recorded and reported symptoms in their children using a validated scoring manual developed by Taylor and colleagues and collected a nasopharyngeal sample on day 2 of any episode [35]. The probe was sent for analysis using semi-quantitative RT-PCR (Allplex Respiratory Full Panel Assay Seegene, South Korea) and the following pathogens were tested: rhinovirus, adenovirus, enteroviruses, respiratory syncytial virus, influenza and coronaviruses (229E, OC43 and NL63). The methodological quality according to Jadad was with 4 points again high [30].

Overall, 57 and 72 samples were positively tested for mRNA from any respiratory virus during prevention with Echinacea and control, respectively (p=0.0074, chi-square test). Similar to results obtained from adults, Echinacea significantly reduced the incidence of enveloped virus infections in children with a total of N=29 samples compared to control with N=47 yielding a significant odds ratio of OR=0.43 (p=0.0038, chi-square test). Again, information relating to coronavirus subtypes was retrieved from study authors: coronavirus incidence was with 13 and 11 positive samples in the intention-to-treat sample insignificantly different between interventions (p>0.1). Two CoV episodes in each group preexisted at inclusion visit and were not regarded for prevention analysis but for evaluation of treatment benefits. In contrast to the earlier study by Jawad et al., here, Ct-values were determined for respective incidence tests, estimating the number of RNA copies as a measure for virus concentration in the nasopharyngeal sample. The higher the Ct-value, the lower is the viral load represented by the sample’s RNA level. Echinacea significantly increased the average Ct-value by 6.1 Ct units from 25.0 [95% CI: 20.5; 29.5] to 31.1 [95% CI: 26.3; 35.9] indicating a strong, -1.81 log or 98.5 % reduction in coronavirus concentration (p=0.0479, Wilcoxon test) relative to Vitamin C. Similar ΔmCt values of 5.53 and 11.92 were observed for variants NL63 and OC43, equaling a virus log reduction by -1.67 and -3.59 (Figure 3), corresponding to an absolute virus reduction of 97.8% and 99.97%.

**Figure 3:**
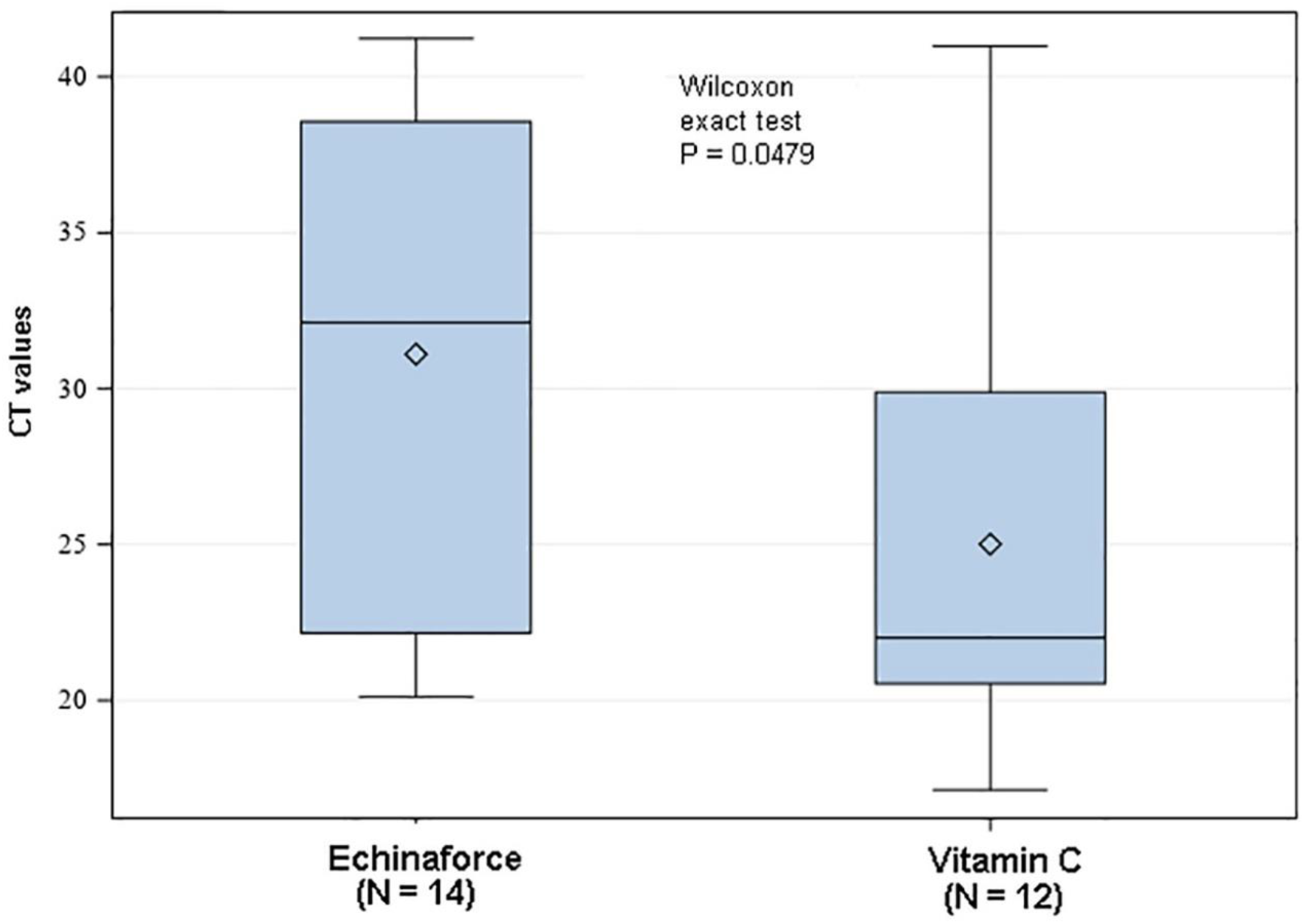
Preventive treatment of children reduced nasopharyngeal coronavirus concentrations significantly by 98.5% in comparison to control (Vitamin C, p<0.05).

### 3.1. Figures, Tables and Schemes

Next, we analyzed the symptomatic development of confirmed coronavirus infections in children treated with Echinacea or control. To this end, an aggregated total symptom sum score was composed of ‘runny nose’, ‘blocked nose’, ‘sneezing’, ‘headache and aching limbs’, ‘sore throat’, ‘cough’, ‘chilliness’, ‘disturbed sleep quality of the child’, ‘malaise’, ‘need for additional care-giving’, each rated as « absent », « mild », « moderate », « severe » or « severity not assessable». The area-under-curve (AUC) indicated a strong, 64.2% reduction from 75.8 [95% CI: 39.8–111.7] to 27.1 [95% CI: 14.8–39.4] score points in children supplemented with Echinacea (p=0.0036). Results are provided for the intention-to-treat (ITT) collective but did not relevantly change in the per protocol group (PP). Apparently, coronavirus infections during Echinaforce prevention were significantly less severe from the very first day of episode until the end of 10 days assessment.

## 4. Discussion

Echinacea owns a long tradition for the prevention and the acute treatment of respiratory tract infections and recent investigations attributed antiviral, immune-modulatory and anti-inflammatory pharmacological actions to the medicinal plant [20,26,36–38]. A wide series of respiratory viruses were shown to be sensitive to lipophilic extracts of freshly harvested *Echinacea purpurea* and an obvious specificity towards enveloped pathogens could be verified [39]. The extract appears to block interaction of viral docking receptors (e.g. hemagglutinin on influenza) with structures on the target cell and is thus expected to prevent infection, although the detailed mechanism of action still has to be elucidated [37]. Antiviral effects on endemic coronaviruses causing typical cold symptoms (CoV-229E) and on highly pathogenic SARS-CoV-1 and MERS-CoV type have been studied in vitro for several years until very recently. These effects could also be confirmed for the newly occurring SARS-CoV-2 [19]. Signer et al. concluded a generally applicable principle of action against coronaviruses, but pointed out that clinical data are needed to confirm their in vitro findings. This work aimed to gather any evidence from in vivo activity against coronavirus infections from already published clinical studies.

Despite the large number of Echinacea clinical trials we identified only two clinical studies that collected nasopharyngeal samples for virus testing [31,34]. Both studies accurately described blinding, randomization, were appropriately controlled and referred to data set from almost 1000 patients treated over a period of 4 months with either tincture or tablets from the same 65% (v/v) ethanolic extract from *Echinacea purpurea*. Due to the low number of included studies, we abstained from pooling results, but to qualitatively describe outcomes from the studies.

Preventive antiviral effects for Echinacea for coronavirus infections were seen in both studies by varying parameters. 4-months supplementation with Echinacea in adults and children reduced the incidence of coronaviruses as part of its effect on enveloped virus infections and virus concentration in nasopharynx in the latter, respectively. Jawad did not measure virus concentrations (Ct-value), and this information was therefore lacking. Ogal employed a newer, more sensitive method for detection that was able to detect as few as 100 viral genome copies. We presume that Echinacea’s antiviral effects generally reduced the amount of viruses, that in Jawad study suppressed below detection limits resulting in decreased incidence with Echinacea and in Ogal et al. resulted in lower Ct-values as even residual, subclinical viral concentrations could still be identified.

Interestingly, prevention with Echinacea reduced the symptomatic development only in children rather than in adults. Jawad et al. did not sample continuously but only upon symptomatic infections and might have missed potential subclinical infections with verum for above reasons. Thereby a reduced incidence (as with Echinacea) would automatically result in missed episodes with no or low symptomatology and skew any evaluation at this level. In contrast, the low virus detection limit in Ogal et al. might have identified more milder/subclinical episodes which would have further reduced symptoms with Echinacea treatment in adults as observed for children. With this respect, incidence (detections) and symptomatology / Ct values must be considered as complementary parameters both indicative for antiviral effects of Echinacea. Taken this into account, we conclude that preventive treatment with Echinacea provides beneficial effects to coronavirus infections in both, adults and children. In the prior infection could be prevented whereas children demonstrated significantly reduced virus loads, symptom reduction as well as shortened duration of illnesses. In both cases, preventive effects of Echinacea against coronaviruses could be ascertained in a controlled clinical setting.

Neither of the referenced studies detected the newly occurring SARS-CoV-2 but only endemic coronavirus strains, as both were carried out prior to 2019. Conclusions pertaining to Covid-19 must be carefully considered but are nevertheless important, since many people rely on immune support by herbal preparations these days and it is important to give an estimation on evidence.

All coronaviruses share structural similarities, like the feature of an enveloped membrane embracing the genetic material, i.e. RNA and spike proteins for attachment to target cells. Echinacea on the other side broadly inhibits enveloped respiratory viruses at physiological concentrations in vitro and in vivo, whereas the exact mode-of-action remains to be elucidated. Due to this broad range of activity, deactivating all measured coronavirus types, including alpha(CoV-229E) and beta-strains (MERS-CoV, SARS-CoV-1, SARS-CoV-2) a generally applicable inhibition can realistically be assumed and the current results of this study point towards clinical relevance of in vitro findings. Though not always reaching statistical significance for the particular virus, the effects of Echinacea against coronaviruses are mirrored and overarched by the effects seen against membranous viruses.

In addition, a meanwhile completed additional clinical study was identified on clinicaltrials.gov (ID: NCT05002179), investigating an *Echinacea purpurea* preparation from fresh herbs in dosages of 2,400 mg – 4,000 mg/d extract per day over 5 months in comparison with non-treatment. The same preparation as in Ogal et al. was used but at the recommended posology for adults. The study was carried out in N=120 adults from November 2020 until May 2021 and routinely collected naso-/ oropharyngeal / blood samples for detection of respiratory virus infections, including SARS-CoV-2. Results suggest a significantly reduced risk for SARS-CoV-2 infections, measured either as symptomatic Covid-19 illness, RT-PCR positive sample or by seroconversion. Summarized over all phases of prevention, 21 and 29 samples tested positive for any virus in the *Echinacea* and control group, of which 5 and 14 samples tested SARS-CoV-2 positive (RR=0.37, Chi square test, p=0.03). Overall, 10 and 14 symptomatic episodes occurred, of which 5 and 8 were COVID-19 (RR=0.70, Chi-square test, p>0.05). EF treatment when applied during acute episodes significantly reduced the overall virus load by at least 2.12 log10 or approx. 99% (t-test, p<0.05), the time to virus clearance by 8.0 days for all viruses (Wilcoxon test, p=0.02) and by 4.8 days for SARS-CoV-2 (p>0.05) in comparison to control. Finally, *Echinacea* treatment significantly reduced fever days (1 day vs. 11 days, Chi square test, p=0.003) [40].

Findings still await publication in peer-reviewed journal and need to be treated with appropriate caution. They were nevertheless mentioned in the current review for the sake of completeness and because they seem to confirm applicability of antiviral benefits of Echinacea to a broad range of coronaviruses, including SARS-CoV-2.

Despite limitations associated with this review (e.g. low number of studies) we believe that our findings are highly relevant as they provide a rather consistent picture on antiviral and preventive benefits of Echinacea in coronavirus infections overall. Also, they provide important implications for the preventive use of Echinacea during Covid-19 epidemic. The reduction of coronavirus loads is medicinally highly relevant in whereas virus concentrations appear to correlate with community transmission, influence illness severity and progression in adults and children [41–44]. Two clinical studies have shown over 98.5% reduction of coronavirus concentration in nasal secretions obtained from children and adults treated with Echinacea. Evidence is still missing that vaccines significantly reduce viral loads, especially of SARS-CoV-2 delta variant and this asset would be a decisive argument for use of Echinacea during Covid-19 pandemic.

Notably, all cited studies applied the same *Echinacea purpurea* extract (Echinaforce®) either as diluted tincture or as tablets, both of which were kept in mouth for a while prior to swallowing. As already hypothesized by Signer et al., pharyngeal administration of the extract may be key to inactivate respiratory viruses at the main entry site prior to infection [19]. Further could any viral attenuation in this region prevent further dissemination of nasal infections to the lungs, as seen with severe progressions of Covid-19. Indeed, a recent meta-analysis found a significant prevention of pneumonia secondary to viral respiratory tract infections upon 2 to 4 months Echinacea prevention [45].

## 5. Conclusions

*Echinacea purpurea* (L.) Moench exhibits direct antiviral activity against a broad range of respiratory pathogens, including coronaviruses. Extracts support the tonic production of interferon and modulate inflammatory cytokines (TNF-a). Nowadays, clinical evidence exists to show how enveloped viruses are effectively prevented in adults and children and this review extrapolates efficacy to coronaviruses as well. Yet unpublished clinical results on SARS-CoV-2 and in vitro experiments provide a good rational for general preventive effects against coronavirus infections.

## Data Availability

All data produced in the present work are contained in the manuscript.

## Author Contributions

Conceptualization SN and GG; formal analysis, KP; writing—original draft preparation, SN; writing—review and editing SR, KAW, SN, OM; All authors have read and agreed to the published version of the manuscript.

## Acknowledgments

We would like to thank Roland Schoop from the A.Vogel AG company for providing us with study details on coronavirus infections.

## Appendix 1

The search strategy and results are detailed in Appendix 1.

