## Appendix 1 for "*Echinacea* as a Potential Force against Coronavirus Infections? A Mini-Review of Randomized Controlled Trials in Adults and Children": Echinacea_Minireview_Appendix1.docx

Study Title

Echinacea as a Potential Force against Coronavirus Infections? A Mini-Review of Randomized Controlled Trials in Adults and Children

Aim

The goal of this review was to identify coronavirus infections in clinical studies on Echinacea and to evaluate preventive and treatment benefits.

Methods

The Preferred Reporting Items for Systematic Reviews and Meta-Analyses (PRISMA) guidelines were followed to conduct and write this review [1].

Eligibility Criteria for Study Selection

Eligible studies were selected based on the following criteria:

(i) studies reporting original data on effect of Echinacea on respiratory tract pathogens
(ii) studies were analyzed if they reported human sampling and assessed the effect of prevention or therapy with *Echinacea species* such as *Echinacea* or *Echinacea purpurea* or *Echinacea* or coneflower, which have been identified as playing a role in evidence for in vivo and in vitro activity against coronavirus infections
(iii) we included randomized controlled trials (RCTs), clinical studies, which assess the effect of Echinacea on respiratory tract pathogens in humans. Abstracts and unpublished studies were not included. When articles contained potentially relevant data not fully reported, the authors were contacted by email for further information.

Outcomes measures

Data extraction was undertaken using a pre-defined form retrieving the following information: primary author’s name, publication date, country, study design, sample size, researched plant species and preparation, duration of treatment (prevention vs acute treatment), dosage, incidence of coronavirus virus infections and virus concentration (Ct-values), information regarding symptomatic development (e.g. total symptom score, episode duration, area-under-curve or similar). The methodological quality of included studies was assessed using the criteria as proposed by Jadad [2].

Search Strategy

We used PubMed and EMBASE databases for the literature search using a combination of keywords, which includes Medical Subject Headings (MeSH)/*Emtree* terms (Embase Subject Headings). The search strategy was limited to English, humans and included all synonyms of the following MeSH terms: Echinacea species OR inclusion coronavirus virus infections, AND Roter Sonnenhut or Purple Coneflower OR Echinacea purpurea OR human Clinical Studies OR Respiratory Tract Infections AND terms related to study design (RCTs). The following search headings/ MeSH terms were used:

(Randomized Controlled Trials as Topic / OR randomized controlled trial/ OR Random Allocation/ OR Double Blind Method/ OR Single Blind Method/ OR clinical trial/ OR clinical trial, phase OR clinical trial, phase ii.pt. OR clinical trial, phase iii.pt. OR clinical trial, clinical studies OR phase iv.pt. OR controlled clinical trial.pt. OR randomized controlled trial.pt. OR multicenter study OR Placebo OR randomly allocated.)

AND (“Echinacea species” (MeSH Terms) OR “Echinacea species” [All Fields]) OR (“Purple Coneflower” (MeSH Terms) OR “Purple Coneflower” [All Fields]) OR (“Echinacea purpurea	” (MeSH Terms) OR “Echinacea purpurea	” [All Fields]) OR (“Echinacea [All Fields] AND “purpurea [All Fields]) OR (“Echinacea species, Roter sonnenhut ” (All Fields) OR (“Echinacea species”(All Fields) AND “Roter”(All Fields) AND “sonnenhut”(All Fields))

AND ("clinical study"[Publication Type] OR "clinical studies as topic"[MeSH Terms] OR "clinical studies"[All Fields]) AND ("respiratory tract infections"[MeSH Terms] OR ("respiratory"[All Fields] AND "tract"[All Fields] AND "infections"[All Fields]) OR "respiratory tract infections"[All Fields]) AND ("human s"[All Fields] OR "humans"[MeSH Terms] OR "humans"[All Fields] OR "human"[All Fields]) ("respiratory system"[MeSH Terms] OR ("respiratory"[All Fields] AND "system"[All Fields]) OR "respiratory system"[All Fields] OR ("respiratory"[All Fields] AND "tract"[All Fields]) OR "respiratory tract"[All Fields]) AND ("virology"[MeSH Subheading] OR "virology"[All Fields] OR "viruses"[All Fields] OR "viruses"[MeSH Terms] OR "virus s"[All Fields] OR "viruse"[All Fields] OR "virus"[All Fields]) AND ("body fluids"[MeSH Terms] OR ("body"[All Fields] AND "fluids"[All Fields]) OR "body fluids"[All Fields])

AND “Respiratory Tract Infections” (MeSH Terms) OR (“Respiratory” [All Fields] AND “Tract [All Fields] AND “Infections” [All Fields]) OR “Respiratory Tract Infections” {All Fields] OR (“Respiratory” [All Fields} AND “Tract” [All Fields] AND “Infections” [All Fields]) OR “Respiratory Tract Infections” [All Fields]) “safety and efficacy ” (MeSH Terms) OR (“safety” [All Fields] AND “efficacy” [MeSH Terms] AND “common cold” (MeSH Terms) OR (“common” [All Fields] AND “cold” [MeSH Terms] OR "disease"[All Fields] OR "diseases"[All Fields] OR "disease s"[All Fields] OR "diseased"[All Fields]) “corona virus” (MeSH Terms) OR (“corona” [All Fields] AND “Virus” [All Fields]) [2] [‘SARS-CoV2)OR ‘SARS’ OR ‘CoV2’] AND [children OR paediatric] AND [adults].

Data Extraction

After applying the eligibility criteria, articles that were relevant for full-text screening were chosen. Two authors worked independently on article screening and data extraction. If there were any disagreements, they were settled by discussion or consensus with a third independent reviewer. The articles were initially screened on the basis of their title, followed by the abstract of the article. The title and abstract of the articles were irrelevant to the present investigation; these were excluded from the secondary screening. The selected articles from the initial screening were assessed for full-text screening to find out the eligibility criteria of the current review.

Results

A total of 1687 articles were identified through database searching including PubMed and EMBASE, of which 988 articles were excluded at the initial screening due to duplicated and irrelevant articles. 60 potentially relevant articles were selected for the full text assessment, of which 58 were further excluded due to studies that were not reported respiratory tract infections, coronavirus, Echinacea species, non-RCT and human studies (n= 57) and combination of Zingiber officinale and Echinacea (n= 1), which was detailed in PRISMA flow chart (Figure 1). Finally, 2 articles were selected for the narrative review, which met the eligibility criteria of the current study.

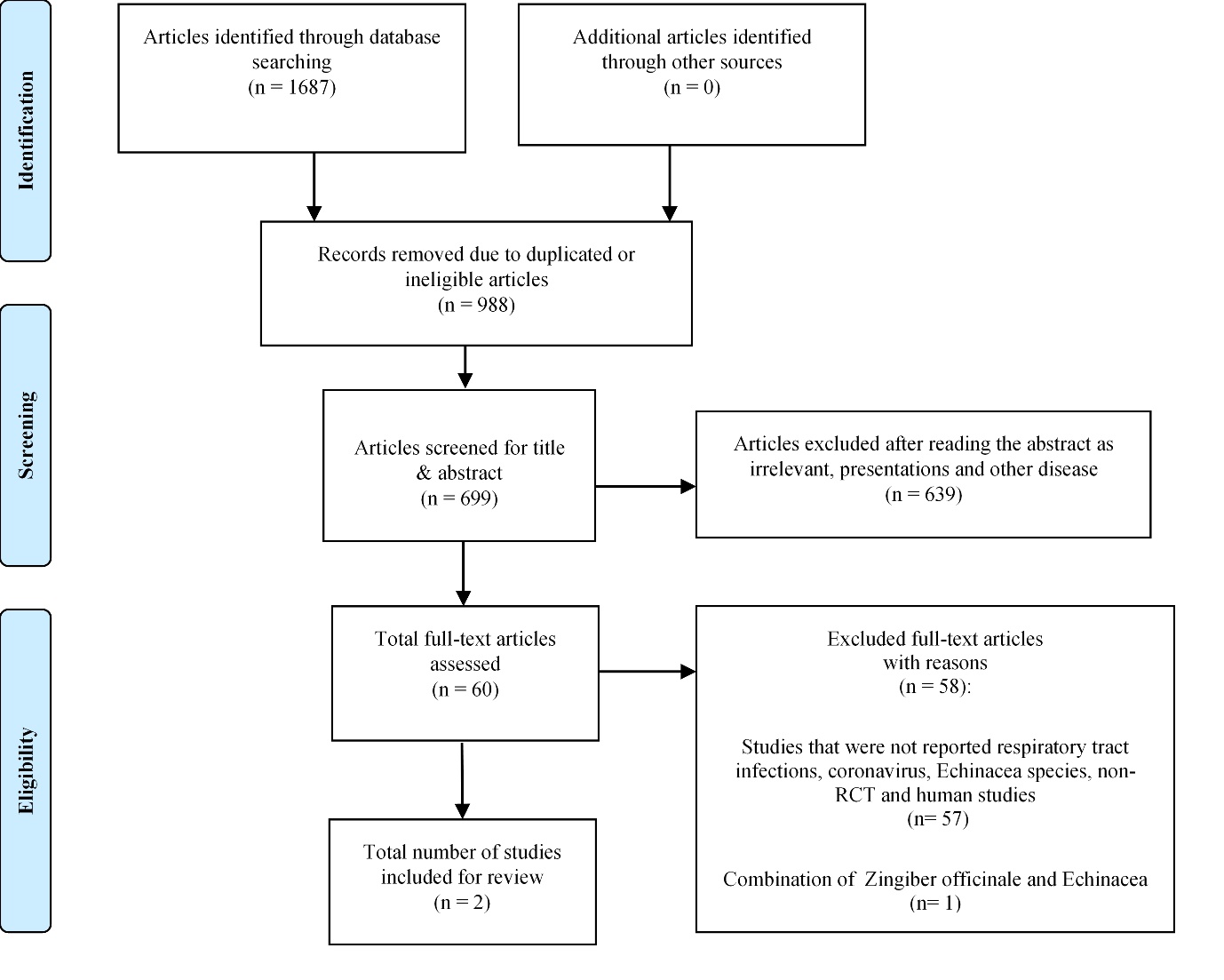

Fig.1: PRISMA flow chart for systematic reviews detailing the database searches, the number of publications screened and eligible results retrieved.

**Annex**

A1. Eligibility criteria

A2. Search strategy

A3. Database specific strategies

A4. Search strategy

A5. Included and excluded articles with justification

A6. Data extraction template

A1 Eligibility criteria.

Table A1 Eligibility criteria.

| **Inclusion criteria** | **Exclusion criteria** |
| --- | --- |
| Randomized controlled trials  Studies that reported the safety and efficacy of Echinacea species against respiratory tract pathogens  Studies that reported the safety and efficacy of Echinacea species against coronavirus  Children and adults population | Conference proceedings, editorials letters and other study designs (Non-RCT).  Studies that were used other than Echinacea species  Studies that did not evaluated the respiratory tract pathogens especially corona virus |

A2 Search strategy

**Table A2 Keywords included in the search strategy for all PubMed and Embase databases; terms searched for in the title and abstract of papers.**

| **Condition** | Echinacea **OR** Echinacea purpurea **OR** Roter Sonnenhut **OR** Purple Coneflower  **AND**  “Respiratory Tract Infections” **OR** “common cold ” **OR** “Respiratory Tract Pathogens” **OR** “coronavirus” |
| --- | --- |
|  | **AND** |
| **Study type term** | Randomized Controlled Trials or Double-blind or Single-blind or Random allocation or Clinical trial or Clinical studies  **AND**  Humans **OR** Adults **OR** Children |

A3 Database specific search strategies

**A3.1 PubMed Search Strategy – limit to human,**

((Randomized Controlled Trials as Topic / OR randomized controlled trial/ OR Random Allocation/ OR Double Blind Method/ OR Single Blind Method/ OR clinical trial/ OR clinical trial, phase OR clinical trial, phase ii.pt. OR clinical trial, phase iii.pt. OR clinical trial, phase iv.pt. OR controlled clinical trial.pt. OR randomized controlled trial.pt. OR multicenter study OR Placebo OR randomly allocated.)

AND (“Echinacea species” (MeSH Terms) OR “Echinacea species” [All Fields]) OR (“Purple Coneflower” (MeSH Terms) OR “Purple Coneflower” [All Fields]) OR (“Echinacea purpurea	” (MeSH Terms) OR “Echinacea purpurea	” [All Fields]) OR (“Echinacea [All Fields] AND “purpurea [All Fields]) OR (“Echinacea species, Roter sonnenhut ” (All Fields) OR (“Echinacea species”(All Fields) AND “Roter”(All Fields) AND “sonnenhut”(All Fields)) AND ("clinical study"[Publication Type] OR "clinical studies as topic"[MeSH Terms] OR "clinical studies"[All Fields]) AND ("respiratory tract infections"[MeSH Terms]

AND “Respiratory Tract Infections” (MeSH Terms) OR (“Respiratory” [All Fields] AND “Tract [All Fields] AND “Infections” [All Fields]) OR “Respiratory Tract Infections” {All Fields] OR (“Respiratory” [All Fields} AND “Tract” [All Fields] AND “Infections” [All Fields]) OR “Respiratory Tract Infections” [All Fields]) “safety and efficacy ” (MeSH Terms) OR (“safety” [All Fields] AND “efficacy” [MeSH Terms] AND “common cold” (MeSH Terms) OR (“common” [All Fields] AND “cold” [MeSH Terms] OR "disease"[All Fields] OR "diseases"[All Fields] OR "disease s"[All Fields] OR "diseased"[All Fields]) “corona virus” (MeSH Terms) OR (“corona” [All Fields] AND “Virus” [All Fields]) [2] [‘SARS-CoV2)OR ‘SARS’ OR ‘CoV2’]

**A3.2. EMBASE Search Strategy – limit to human,**

Echinacea species’/exp OR ‘Echinacea species’ :ab,ti OR ‘Echinacea purpurea	’ /exp OR ‘Echinacea purpurea	’: ab, ti OR ‘Echinacea purpurea	’/exp OR ‘Purple Coneflower’/ exp OR ‘Purple Coneflower’ :ab,ti OR ‘Purple Coneflower’ / exp OR ‘Purple Coneflower’ / exp OR exp Echinacea species / OR ‘Roter sonnenhut’ OR Echinacea purpurea.mp OR Purple Coneflower.mp OR Purple Coneflower.mp OR PC.mp AND (Respiratory Tract Infections /exp OR common, cold.mp or common cold /exp OR Respiratory Tract * Infections. ti,ab. OR common cold. ti,ab OR Respiratory Tract Infections.mp. or exp Respiratory Tract Infections / OR common cold.mo. or exp common cold /) AND (Incidence.mp. or exp safety/ OR efficacy.mp or exp efficacy OR epidemiological data/ OR disease progression/ OR disease activity/ OR ‘Randomized Controlled Trials *.tw’ OR ‘Random Allocation.tw’ OR ‘Double Blind’ OR ‘Randomized Controlled Trials.tw’ OR ‘clinical studies’ OR ‘clinical trial.tw’ or Placebo Controlled Trials /) AND Limit to (english language, human AND conference abstract AND conference paper AND conference proceeding AND “conference review”) AND yr=”

A4 Search results

**PubMed- Search results**

| # | **PubMed** |  |
| --- | --- | --- |
| **Search** | **Keyword** | **Results** |
| 1 | Tier 1: Echinacea Purpurea | 1,331 |
| 2 | ((Echinacea) OR (Roter sonnenhut)) OR (Purple cone flower) | 1,340 |
| 3 | Echinacea | 1,338 |
| 4 | Roter sonnenhut | 0 |
| 5 | Purple cone flower | 8 |
| 6 | (Echinacea Purpurea) AND (Respiratory Tract Infections) | 203 |
| 7 | ((Echinacea Purpurea) AND (Respiratory Tract Infections)) AND (Randomised controlled trail) | 0 |
| 8 | (((Respiratory tract infections) ) OR (common cold)) AND (Echinacea) | 231 |
| 9 | (Respiratory Tract Infections) AND (Echinacea) | 203 |
| 10 | (Echinacea) AND (common cold) | 155 |
| 11 | (((safety and efficacy)) AND (Respiratory Tract Infections)) AND (Echinacea) | 19 |
| 12 | (Echinacea) AND (safety and efficacy) | 65 |
| 13 | (Randomized controlled trail) AND (Echinacea) | 0 |
| 14 | (((Randomized controlled trail) OR (Double blind)) OR (single blind)) OR (clinical trial) | 1,308,606 |
| 15 | (Randomized Controlled Trial) AND (Safety and efficacy) | 53,404 |
| 16 | (((Randomized Controlled Trial) OR (single blind)) AND (clinical trial)) AND (Echinacea Purpurea) | 109 |
| 17 | ((((((((Randomized controlled trail) OR (Double blind)) ) OR (single blind)) OR (clinical trial) ) AND (Echinacea)) OR (Roter sonnenhut)) OR (Purple cone flower) | 194 |
| 18 | (Randomized controlled trial) AND (Respiratory Tract Infections) | 14,460 |
| 19 | Respiratory tract infections | 493,848 |
| 20 | ((Clinical studies) AND (respiratory tract infections)) AND (humans) | 31,002 |
| 21 | (((Respiratory tract infections) AND (humans)) AND (Echinacea)) AND (clinical studies) | 83 |
| 22 | (Respiratory tract viruses) AND (body fluids) | 1,279 |
| 23 | ((Echinacea) AND (respiratory tract viruses)) AND (body fluids) | 0 |
| 24 | (((respiratory tract viruses) AND (body fluids)) OR (Roter sonnenhut)) OR (Purple cone flower) | 1,287 |
| 25 | ((Randomized controlled trial) AND (Echinacea)) AND (Respiratory Tract Infections) | 57 |

**Embase- Search results**

| # | **EMBASE (2011-2021)** |  |
| --- | --- | --- |
| **Search** | **Keyword** | **Results** |
| 1 | Tier 1: Echinacea Purpurea $ | 1,338 |
| 2 | Echinacea OR Roter sonnenhut $ OR Purple cone flower | 1,350 |
| 3 | Echinacea $ | 1,344 |
| 4 | Roter sonnenhut $ | 0 |
| 5 | Purple cone flower $ | 11 |
| 6 | Echinacea Purpurea AND Respiratory Tract Infections $ | 218 |
| 7 | Echinacea Purpurea $ AND Respiratory Tract Infections AND Randomised controlled trail $ | 0 |
| 8 | Respiratory tract infections OR common cold $ AND Echinacea | 229 |
| 9 | Respiratory Tract Infections $ AND Echinacea | 218 |
| 10 | Echinacea $ AND common cold | 162 |
| 11 | safety and efficacy $ AND Respiratory Tract Infections AND Echinacea $ | 18 |
| 12 | Echinacea AND Safety and efficacy $ | 69 |
| 13 | Randomized controlled trail $ AND Echinacea | 0 |
| 14 | Randomized controlled trail OR Double blind $ OR single blind OR clinical trial $ | 1,313,596 |
| 15 | Randomized Controlled Trial $ AND Safety and efficacy | 54,398 |
| 16 | Randomized Controlled Trial OR single blind $ AND clinical trial AND Echinacea Purpurea $ | 112 |
| 17 | Randomized controlled trail $ OR Double blind $ OR single blind OR clinical trial $ AND Echinacea OR Roter sonnenhut $ OR Purple cone flower | 198 |
| 18 | Randomized controlled trial AND Respiratory Tract Infections | 13,260 |
| 19 | Respiratory tract infections $ | 494,851 |
| 20 | Clinical studies $ AND respiratory tract infections AND humans $ | 31,212 |
| 21 | Respiratory tract infections AND humans $ AND Echinacea AND clinical studies$ | 87 |
| 22 | Respiratory tract viruses $ AND body fluids | 1,286 |
| 23 | Echinacea AND respiratory tract viruses $ AND body fluids | 0 |
| 24 | Respiratory tract viruses AND body fluids$ OR Roter sonnenhut $ OR Purple cone flower | 1,278 |
| 25 | Randomized controlled trial AND Echinacea$ AND Respiratory Tract Infections | 59 |

A5: Included and excluded articles with justification

| **S.NO** | **Author** | **Year** | **Title** | **Reason for exclusion and inclusion** | **Links** |
| --- | --- | --- | --- | --- | --- |
| 1 | Motahareh Boozari et al | 2021 | Natural products for Corona virus infection prevention and treatment regarding to previous coronavirus infections and novel studies. | Excluded because NO evidence of randomised control trail | <https://pubmed.ncbi.nlm.nih.gov/32985017/> |
| 2 | Tibebeselassie Seyoum Keflie et al | 2021 | Micronutrients and bioactive substances: Their potential roles in combating Corona virus infection | Excluded because NO evidence of randomised control trail | https://pubmed.ncbi.nlm.nih.gov/33450678/ |
| 3 | Andreas Hensel et al | 2020 | Challenges at the Time of Corona virus infection: Opportunities and Innovations in Antivirals from Nature | Excluded because NO evidence of randomised control trail | https://pubmed.ncbi.nlm.nih.gov/32434254/ |
| 4 | Johanna Signer et al | 2020 | In vitro virucidal activity of Echinaforce®, an Echinacea purpurea preparation, against coronaviruses, including common cold coronavirus 229E and SARS-CoV-2 | Excluded because NO evidence of randomised control trail | https://pubmed.ncbi.nlm.nih.gov/32907596/ |
| 5 | Shaden A M Khalifa et al | 2021 | Screening for natural and derived bio-active compounds in preclinical and clinical studies: One of the frontlines of fighting the coronaviruses pandemic | Excluded because NO evidence of randomised control trail | https://pubmed.ncbi.nlm.nih.gov/33067112/ |
| 6 | M F Nagoor Meeran et al | 2021 | Can *Echinacea* be a potential candidate to target immunity, inflammation, and infection - The trinity of coronavirus disease 2019 | Excluded because NO evidence of randomised control trail | https://pubmed.ncbi.nlm.nih.gov/33585706/ |
| 7 | Shiv Bharadwaj et al | 2021 | Structure-Based Identification of Natural Products as SARS-CoV-2 M pro Antagonist from *Echinacea angustifolia* Using Computational Approaches | Excluded because NO evidence of randomised control trail | https://pubmed.ncbi.nlm.nih.gov/33672054/ |
| 8 | Marlies Karsch-Völk et al | 2014 | Echinacea for preventing and treating the common cold | Excluded because NO evidence of Corona virus infection | https://pubmed.ncbi.nlm.nih.gov/24554461/ |
| 9 | Sholto David et al | 2019 | Echinacea for the prevention and treatment of upper respiratory tract infections: A systematic review and meta-analysis | Excluded because NO evidence of Corona virus infection | https://pubmed.ncbi.nlm.nih.gov/31126553/ |
| 10 | Giulia Dante et al | 2014 | Herbal therapies in pregnancy: what works? | Excluded because NO evidence of Corona virus infection | https://pubmed.ncbi.nlm.nih.gov/24535321/ |
| 11 | Songie Choi et al | 2017 | A systematic review of the pharmacokinetic and pharmacodynamic interactions of herbal medicine with warfarin | Excluded because NO evidence of Corona virus infection | https://pubmed.ncbi.nlm.nih.gov/28797065/ |
| 12 | Aaron S Griffin et al | 2018 | Alternative therapies for chronic rhinosinusitis: A review | Excluded because NO evidence of Corona virus infection | https://pubmed.ncbi.nlm.nih.gov/29554408/ |
| 13 | Attila Oláh et al | 2017 | Echinacea purpurea-derived alkylamides exhibit potent anti-inflammatory effects and alleviate clinical symptoms of atopic eczema | Excluded because NO evidence of Corona virus infection | https://pubmed.ncbi.nlm.nih.gov/28610718/ |
| 14 | Philippe Vorilhon et al | 2019 | Efficacy of vitamin C for the prevention and treatment of upper respiratory tract infection. A meta-analysis in children | Excluded because NO evidence of Corona virus infection | https://pubmed.ncbi.nlm.nih.gov/30465062/ |
| 15 | Ramon Weishaupt et al | 2020 | Safety and Dose-Dependent Effects of Echinacea for the Treatment of Acute Cold Episodes in Children: A Multicenter, Randomized, Open-Label Clinical Trial | Excluded because NO evidence of Corona virus infection | <https://pubmed.ncbi.nlm.nih.gov/33333722/> |
| 16 | Andreas Schapowal et al | 2015 | Echinacea reduces the risk of recurrent respiratory tract infections and complications: a meta-analysis of randomized controlled trials | Excluded because NO evidence of Corona virus infection | https://pubmed.ncbi.nlm.nih.gov/25784510/ |
| 17 | Mehdi Safarabadi et al | 2017 | Comparing the Effect of Echinacea and Chlorhexidine Mouthwash on the Microbial Flora of Intubated Patients Admitted to the Intensive Care Unit | Excluded because NO evidence of Corona virus infection | https://pubmed.ncbi.nlm.nih.gov/29184589/ |
| 18 | M Jawad et al | 2012 | Safety and Efficacy Profile of Echinacea purpurea to Prevent Common Cold Episodes: A Randomized, Double-Blind, Placebo-Controlled Trial | Included | https://pubmed.ncbi.nlm.nih.gov/23024696/ |
| 19 | Surabhi Sinha et al | 2015 | Immunomodulators in warts: Unexplored or ineffective? | Excluded because NO evidence of Corona virus infection and Echinacea species | https://pubmed.ncbi.nlm.nih.gov/25814698/ |
| 20 | E Tiralongo et al | 2012 | Randomised, double blind, placebo-controlled trial of echinacea supplementation in air travellers | Excluded because NO evidence of Corona virus infection | https://pubmed.ncbi.nlm.nih.gov/22229040/ |
| 21 | Andreas Schapowal et al | 2013 | Efficacy and safety of Echinaforce in respiratory tract infections | Excluded because NO evidence of Corona virus infection | https://pubmed.ncbi.nlm.nih.gov/23263637/ |
| 22 | F Isbaniah et al | 2011 | Echinacea purpurea along with zinc, selenium and vitamin C to alleviate exacerbations of chronic obstructive pulmonary disease: results from a randomized controlled trial | Excluded because NO evidence of Corona virus infection | https://pubmed.ncbi.nlm.nih.gov/21062330/ |
| 23 | Mehdi Mesri et al | 2021 | The effects of combination of Zingiber officinale and Echinacea on alleviation of clinical symptoms and hospitalization rate of suspected Corona virus infection outpatients: a randomized controlled trial | Excluded due to combination of Zingiber officinale and Echinacea and no evidence of respiratory tract infections | <https://pubmed.ncbi.nlm.nih.gov/33787192/> |
| 24 | József Haller et al | 2013 | The anxiolytic potential and psychotropic side effects of an echinacea preparation in laboratory animals and healthy volunteers | Excluded because NO evidence of Respiratory tract infections | <https://pubmed.ncbi.nlm.nih.gov/22451347/> |
| 25 | J Orengo et al | 2012 | Evaluating the efficacy of cinnamaldehyde and Echinacea purpurea plant extract in broilers against Eimeria acervulina | Excluded because NO evidence of Respiratory tract infections | https://pubmed.ncbi.nlm.nih.gov/21996002/ |
| 26 | Osama G Abdel-Naby Awad et al | 2019 | Echinacea can help with Azithromycin in prevention of recurrent tonsillitis in children | Excluded because NO evidence of Corona virus infection and randomised control trails | <https://pubmed.ncbi.nlm.nih.gov/32487336/> |
| 27 | Mercedes Ogal et al | 2021 | Echinacea reduces antibiotic usage in children through respiratory tract infection prevention: a randomized, blinded, controlled clinical trial | Included | https://pubmed.ncbi.nlm.nih.gov/33832544/ |
| 28 | Hannah Ayrle et al | 2021 | Effects of an oral hydro-ethanolic purple coneflower extract on performance, clinical health and immune parameters in calves | Excluded because NO evidence of Corona virus infection | https://pubmed.ncbi.nlm.nih.gov/34144282/ |
| 29 | John Grbic et al | 2011 | A phase II trial of a transmucosal herbal patch for the treatment of gingivitis | Excluded because NO evidence of Corona virus infection and Echinacea species | https://pubmed.ncbi.nlm.nih.gov/21965490/ |
| 30 | S Dabbou et al | 2016 | Rabbit dietary supplementation with pale purple coneflower. 1. Effects on the reproductive performance and immune parameters of does | Excluded because NO evidence of Corona virus infection and human study | https://pubmed.ncbi.nlm.nih.gov/26763800/ |
| 31 | Noah Samuels et al | 2012 | Effect of an herbal mouth rinse in preventing periodontal inflammation in an experimental gingivitis model: a pilot study | Excluded because NO evidence of Corona virus infection and Echinacea species | https://pubmed.ncbi.nlm.nih.gov/22479786/ |
| 32 | D Melchart et al | 2000 | Echinacea for preventing and treating the common cold | Excluded because NO evidence of Corona virus infection | <https://pubmed.ncbi.nlm.nih.gov/10796553/> |
| 33 | Francesca Toselli et al | 2009 | Echinacea metabolism and drug interactions: the case for standardization of a complementary medicine | Excluded because NO evidence of Corona virus infection | https://pubmed.ncbi.nlm.nih.gov/19427870/ |
| 34 | M A Flannery et al | 1999 | From rudbeckia to echinacea: the emergence of the purple cone flower in modern therapeutics | Excluded because NO evidence of Corona virus infection | https://pubmed.ncbi.nlm.nih.gov/11623947/ |
| 35 | Theophilus B Kwofie et al | 2012 | Respiratory viruses in children hospitalized for acute lower respiratory tract infection in Ghana | Excluded because NO evidence of Corona virus infection and Echinacea species | https://pubmed.ncbi.nlm.nih.gov/22490115/ |
| 36 | María J Giuffrida et al | 2014 | Increased cytokine/chemokines in serum from asthmatic and non-asthmatic patients with viral respiratory infection | Excluded because NO evidence of Corona virus infection and Echinacea species | https://pubmed.ncbi.nlm.nih.gov/23962134/ |
| 37 | Vinti Goel et al | 2005 | A proprietary extract from the echinacea plant (Echinacea purpurea) enhances systemic immune response during a common cold | Excluded because NO evidence of randomised control trail | https://pubmed.ncbi.nlm.nih.gov/16177972/ |
| 38 | Carty K Y Chan et al | 2020 | Preventing Respiratory Tract Infections by Synbiotic Interventions: A Systematic Review and Meta-Analysis of Randomized Controlled Trials | Excluded because NO evidence of Corona virus infection | https://pubmed.ncbi.nlm.nih.gov/31996911/ |
| 39 | Marina Azambuja Amaral et al | 2017 | Network meta-analysis of probiotics to prevent respiratory infections in children and adolescents | Excluded because NO evidence of Corona virus infection | https://pubmed.ncbi.nlm.nih.gov/28052594/ |
| 40 | Lauren Fontana et al | 2019 | Respiratory Virus Infections of the Stem Cell Transplant Recipient and the Hematologic Malignancy Patient | Excluded because NO evidence of Corona virus infection | https://pubmed.ncbi.nlm.nih.gov/30940462/ |
| 41 | Claire L Hoban et al | 2019 | Analysis of spontaneous adverse drug reactions to echinacea, valerian, black cohosh and ginkgo in Australia from 2000 to 2015 | Excluded because NO evidence of Corona virus infection | https://pubmed.ncbi.nlm.nih.gov/31113761/ |
| 42 | Shaden A M Khalifa et al | 2021 | Screening for natural and derived bio-active compounds in preclinical and clinical studies: One of the frontlines of fighting the coronaviruses pandemic | Excluded because NO evidence of randomised control trail | https://pubmed.ncbi.nlm.nih.gov/33067112/ |
| 43 | Pius S Fasinu et al | 2019 | Herbal Interaction With Chemotherapeutic Drugs-A Focus on Clinically Significant Findings | Excluded because NO evidence of randomised control trail | https://pubmed.ncbi.nlm.nih.gov/31850232/ |
| 44 | Sara M Handy et al | 2021 | HPLC-UV, Metabarcoding and Genome Skims of Botanical Dietary Supplements: A Case Study in Echinacea | Excluded because NO evidence of randomised control trail | https://pubmed.ncbi.nlm.nih.gov/33445185/ |
| 45 | Kathryn M Docherty et al | 2019 | Soil microbial restoration strategies for promoting climate-ready prairie ecosystems | Excluded because NO evidence of Corona virus infection | https://pubmed.ncbi.nlm.nih.gov/30680826/ |
| 46 | Taha Kumosani et al | 2020 | Evaluation in broilers of aerosolized nanoparticles vaccine encapsulating imuno-stimulant and antigens of avian influenza virus/Mycoplasma gallisepticum | Excluded because NO evidence of randomised control trail | https://pubmed.ncbi.nlm.nih.gov/32867774/ |
| 47 | Grzegorz Świderski et al | 2020 | Spectroscopic, Theoretical and Antioxidant Study of 3d-Transition Metals (Co (II), Ni(II), Cu(II), Zn(II) Complexes with Cichoric Acid | Excluded because NO evidence of Corona virus infection | https://pubmed.ncbi.nlm.nih.gov/32664569/ |
| 48 | Na Li et al | 2021 | Enhanced phytoremediation of PAHs and cadmium contaminated soils by a Mycobacterium | Excluded because NO evidence of Corona virus infection | https://pubmed.ncbi.nlm.nih.gov/33254925/ |
| 49 | Dominique Turck et al | 2020 | Anxiofit-1 and reduction of subthreshold and mild anxiety: evaluation of a health claim pursuant to Article 14 of Regulation (EC) No 1924/2006 | Excluded because NO evidence of Corona virus infection | https://pubmed.ncbi.nlm.nih.gov/33117457/ |
| 50 | Ewa Skała et al | 2020 | Caffeoylquinic Acids with Potential Biological Activity from Plant In vitro Cultures as Alternative Sources of Valuable Natural Products | Excluded because NO evidence of Corona virus infection | https://pubmed.ncbi.nlm.nih.gov/32048962/ |
| 51 | Andra Dumitrascu et al | 2019 | Acute respiratory and urinary tract infections in medical practice : a selection of complementary medicines | Excluded because NO evidence of Corona virus infection | https://pubmed.ncbi.nlm.nih.gov/31021574/ |
| 52 | Maria Olga Kokornaczyk et al | 2019 | Phenomenological Characterization of Low-Potency Homeopathic Preparations by Means of Pattern Formation in Evaporating Droplets | Excluded because NO evidence of Corona virus infection | https://pubmed.ncbi.nlm.nih.gov/30625507/ |
| 53 | Shawn D Taylor et al | 2019 | Estimating flowering transition dates from status-based phenological observations: a test of methods | Excluded because NO evidence of Corona virus infection | https://pubmed.ncbi.nlm.nih.gov/31579602/ |
| 54 | Yi Yuan et al | 2021 | Interventions for preventing influenza: An overview of Cochrane systematic reviews and a Bayesian network meta-analysis | Excluded because NO evidence of Corona virus infection | https://pubmed.ncbi.nlm.nih.gov/34544670/ |
| 55 | Liqun Hou et al | 2019 | Study on the efficiency of phytoremediation of soils heavily polluted with PAHs in petroleum-contaminated sites by microorganism | Excluded because NO evidence of Corona virus infection | https://pubmed.ncbi.nlm.nih.gov/31485937/ |
| 56 | Maureen L Page et al | 2019 | Pollinator effectiveness in a composite: a specialist bee pollinates more florets but does not move pollen farther than other visitors | Excluded because NO evidence of Corona virus infection | https://pubmed.ncbi.nlm.nih.gov/31713237/ |
| 57 | Hans-Heinrich Henneicke-von Zepelin et al | 2019 | Non-interventional observational study broadens positive benefit-risk assessment of an immunomodulating herbal remedy in the common cold | Excluded because NO evidence of Corona virus infection | https://pubmed.ncbi.nlm.nih.gov/31074674/ |
| 58 | Esin Akyüz et al | 2019 | Protein-Protected Gold Nanocluster-Based Biosensor for Determining the Prooxidant Activity of Natural Antioxidant Compounds | Excluded because NO evidence of Corona virus infection | https://pubmed.ncbi.nlm.nih.gov/31459484/ |
| 59 | Alfredo Guarino et al | 2013 | Definitions and outcomes of nutritional interventions in children with respiratory infections: the approach of the COMMENT initiative | Excluded because NO evidence of Corona virus infection | https://pubmed.ncbi.nlm.nih.gov/24296796/ |
| 60 | Song Mao et al | 2013 | Vitamin D supplementation and risk of respiratory tract infections: a meta-analysis of randomized controlled trials | Excluded because NO evidence of Corona virus infection | https://pubmed.ncbi.nlm.nih.gov/23815596/ |

A6. Data Extraction Template

| **Source details** | Study code |
| --- | --- |
|  | Author |
|  | Year |
|  | Title |
| **Methodology** | Study design |
|  | Cohort |
| **Participant characteristics** | Sample size |
|  | Ethnicity |
|  | Age |
|  | Gender |
|  | Respiratory tract infection |
|  | Timing of data collection |
|  | Methodology (assessment of) for Respiratory tract pathogens (coronavirus) |
|  | Treatment |
|  | Screening |
| **Outcome (Prevalence & Epidemiology)** | Total symptom score |
|  | Episode duration |
|  | Area-under-curve or similar |
