## Supplementary figures and images for "*Echinacea* as a Potential Force against Coronavirus Infections? A Mini-Review of Randomized Controlled Trials in Adults and Children"

### Graphical_abstract_Echinacea_CoV.jpg

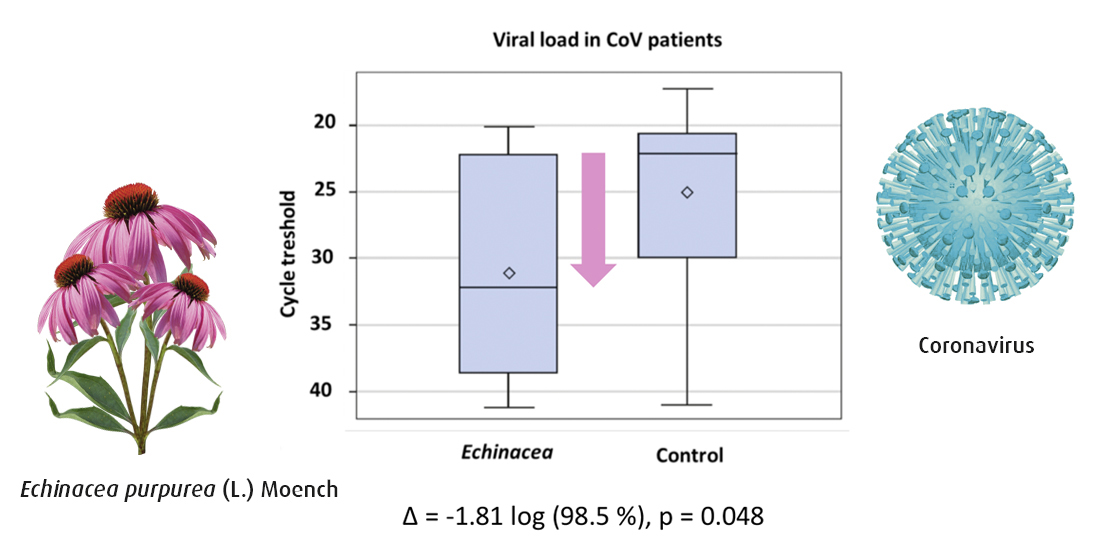
